# Characterization of small fiber neuropathy in hypermobile Ehlers Danlos Syndrome

**DOI:** 10.1101/2022.02.17.22271061

**Authors:** Aurore Fernandez, Bérengère Aubry-Rozier, Mathieu Vautey, Chantal Berna, Marc R. Suter

**Affiliations:** Faculty of biology and Medicine, University of Lausanne, Switzerland; Pain Center, Division of anesthesiology, Lausanne University Hospital (CHUV), Switzerland; Center for integrative and complementary medicine, Division of anesthesiology, (CHUV); Department of Rheumatology, (CHUV)

## Abstract

Hypermobile Ehlers Danlos Syndrome (hEDS)/hypermobility spectrum disorders (HSD) are incapacitating and painful syndromes involving a generalized connective tissue disorder with joint hypermobility and musculoskeletal complications. A neuropathic component is clinically likely given frequent burning sensations, hypoesthesia, or allodynia. Small fiber neuropathy (SFN) refers to the dysfunction or damage of A-δ and C-fibers, which relay thermal and nociceptive information as well as mediating autonomic function. SFN has been suggested by prior studies in hEDS but these early findings (case series N≤20) with sole reliance on intraepidermal nerve fiber density (IENFD) called for a larger sample combined with functional testing.

In this retrospective chart extraction from 79 hEDS/HSD patients referred to a pain center due to neuropathic pain or dysautonomia, both functional (Quantitative Sensory Testing (QST), N=79) and structural (IENFD, N=69) evaluations of small nerve fibers were analyzed in combination with clinical data and standardized questionnaires.

All the patients reported moderate to severe pain interfering with daily life. A decreased thermal detection (QST) was shown in 55/79 patients (70%) and a decreased IENFD in 54/69 patients (78%). Hence a small fiber neuropathy (both abnormal IENFD and QST) was definite in 40/69 patients (58%), possible in 23/69 patients (33%) and excluded in only 6/69 patients (9%).

These results add strong evidence for a peripheral neuropathic contribution to pain symptoms in hEDS/HSD, in addition to the known nociceptive and central sensitization components. Such neuropathic contribution could raise the hypothesis of a neurological cause of hEDS, the only EDS syndrome still without a known genetic cause. Hence, our data is leading the way to a better stratification of this very heterogeneous population, which could improve symptom management and expand pathophysiological research.

## Introduction

Ehlers Danlos syndromes (EDS) regroup a set of clinically and genetically heterogeneous heritable connective tissue disorders [1]. Among the 13 different subtypes of EDS, the hypermobile form (hEDS) is the most frequent and the only one without an identified genetic mutation. The diagnosis of hEDS, according to the revised criteria of 2017, relies on clinical symptoms including generalized joint hypermobility as well as an association of systemic manifestations of connective tissue disorder and musculoskeletal complications [1]. Patients suffering from symptomatic joint hypermobility, yet not fulfilling the criteria for a hEDS, enter the Hypermobility Spectrum Disorder (HSD) category [2].

Associated symptoms such as fatigue [3], dysautonomia, as well as psychiatric and psychological co-morbidities [4] are described in both hEDS and HSD [5]. Chronic pain is frequent in hEDS/HSD with a high impact on daily life [3, 6]. Patients report musculoskeletal pain, but also headaches, gastro-intestinal, genito-urinary and pelvic pain [7-11] and often describe burning sensations, paresthesia and allodynia, suggesting a neuropathic component. Based on a self-administered questionnaire in N=44 EDS patients (29 hEDS and 15 classic EDS), 68% suffered from at least “probable” neuropathic pain [12].

Neuropathic pain is caused by a lesion or a disease of the somatosensory system [13], including the peripheral nerve fibers. One can distinguish large and thickly myelinated Aβ sensory nerve fibers, promoting the conduction of touch, pressure, proprioception and vibration signals from thinly myelinated A-δ and unmyelinated C-fibers sensing temperature, nociceptive inputs and itch and also mediating autonomic functions (sudomotor, thermoregulatory, cardiovascular, gastrointestinal and urogenital) [14]. Small fiber neuropathy (SFN) refers to the dysfunction or damage of A-δ and C-fibers. There is no consensus on the diagnosis of SFN [14-16], which currently relies on a combination of clinical examination, functional testing (such as Quantitative Sensory Testing, QST) and structural quantification of IntraEpidermal Nerve Fiber Density (IENFD) [15, 17].

Small fiber neuropathy has been suggested in hEDS patients in case reports [18, 19] and in a study revealing decreased IENFD in 20 consecutive hEDS patients [20]. These studies provided early findings of small fiber involvement in hEDS, but their limited sample size and sole reliance on IENFD called for further investigations. In this study, we report both structural (IENFD) and functional (QST) evaluations of small nerve fibers in patients suffering from hEDS or HSD. To our knowledge, this study represents the largest cohort to date of hEDS/HSD patients having undergone standardized sensory, histological and psychological assessments.

## Methods

### Subjects

This retrospective chart analysis was approved by the Ethics Committee, Vaud, Switzerland (CERVD 2019-00093). The study population consisted of adult patients suffering from hEDS/HSD followed at the Department of Rheumatology, Lausanne University Hospital, who had signed general informed consent allowing further use of their clinical data. The following three criteria were required for a hEDS diagnosis [1]: (1) Generalized Joint Hypermobility, based on the Beighton score; (2) presence of ≥ 2 of 3 following characteristics: A. ≥ 5 systemic manifestations; B. positive family history; C. musculoskeletal involvement; (3) exclusion of other diagnoses explaining the symptoms. Patients with symptomatic joint hypermobility, yet not fitting these criteria, were diagnosed as Hypermobility Spectrum Disorder (HSD) [2]. Since 2017, hypermobile patients were referred to the Pain Center of Lausanne University Hospital if there was a suspicion of small fiber neuropathy based on anamnestic complaints (hypoesthesia, burning pain, allodynia, dysesthesia or dysautonomia) (Figure 1). Patients were evaluated by a pain physician, filled standardized questionnaires, and underwent QST. Skin biopsies became available from 2020 on and were offered to complete the assessment (hence up to 3 years apart from the initial clinical evaluation). In patients with a suspicion of SFN, an electroneuromyogram (ENMG) was suggested to be performed by neurologists to exclude large fiber involvement.

**Figure 1.**
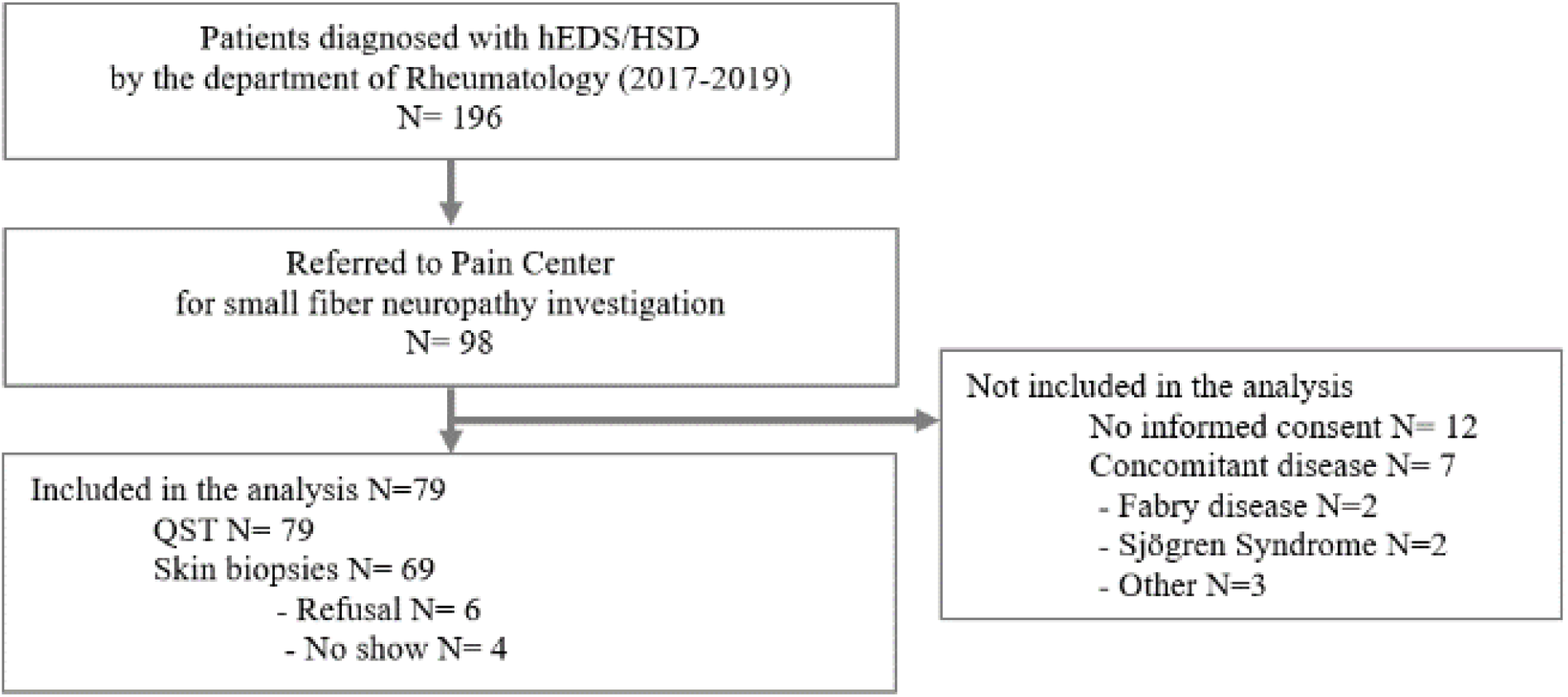
Flowchart illustrating the clinical path and the inclusion of study subjects. hEDS=hypermobile Ehlers Danlos Syndrome. HSD=Hypermobility Spectrum Disorder. QST=Quantitative Sensory Testing

An average age and gender-matched healthy control group of N=23 was recruited prospectively (CERVD 2020-02259). Exclusion criteria were: neurological symptoms and possible causes of neuropathy such as diabetes mellitus, thyroid dysfunction, hypovitaminosis B12, acute infection, nerve entrapment, HIV or cancer. Participants underwent QST and answered questionnaires.

### Symptoms assessment: pain, rheumatological and psychological comorbidities

Pain intensity and interference with daily life were assessed with the Brief Pain Inventory (BPI) [21]. Pain localization was assessed using the paper body map of the BPI and neuropathic characteristics evaluated with the Douleur Neuropathique 4 questionnaire (DN4) [22]. The widespread pain index (WPI) was used to quantify the pain distribution and the symptom severity scale (SSS) to evaluate symptoms frequently associated with widespread pain such as fatigue, unrestful sleep and cognitive symptoms [23]. We evaluated the psychological health using the Hospital Anxiety and Depression Scale (HADS) [24], fear of movement through the TAMPA scale of kinesiophobia (TSK) [25], and the Pain Catastrophizing Scale (PCS) [26]. Quality of life was assessed through the World Health Organization Quality of Life BREF questionnaire (WHO-QOL BREF) [27]. Raw WHO-QOL-BREF scores were converted into the transformed domains scores ranging from 0 to 100 for physical health, psychological domain, social relationships and environment [27]. The Small Fiber Neuropathy and Symptoms Inventory Questionnaire (SFN-SIQ) was collected [28]. The 13-items of this self-assessment tool investigate the two clinical presentations of small fiber neuropathy, i.e. autonomic symptoms (changes in sweating pattern, diarrhea, constipation, urinary tract problems like hesitancy and incontinence, dry eyes, dry mouth, orthostatism, palpitations, hot flushes) and sensory symptoms (sensitive leg skin, burning feet, sheet intolerance, and nocturnal restless legs). Each item is scored on a 4-point Likert scale (0-never present; 1-sometimes, 2-often, and 3-always present). The SFN-SIQ validated cut-off for this scale. The symptom severity of articular and extra-articular manifestations was assessed by a rheumatologist specialized in hEDS/HSD patients, using the Clinical Somatosensory Scale (CSS-16), a 16 items questionnaire assessing pain, fatigue, sleep score is the sum of each item. There is no formally disturbance, mobility, skin, dysautonomia, cardiac, bleeding, gastrointestinal, bladder, temporomandibular joint, ear-nose-throat, lung tract, sexual and cognitive symptoms. Each item was scored on a Likert scale of severity from 0 (absent) to 4 (severe) [5]. The number of participants who answered each of the questionnaires is reported in Table 1.

**Table 1.** Demographic characteristics of the study population and symptoms description. Mean scores, standard deviation and percentage of positivity=N reaching clinically validated cut-off of scores (considered cut-off) are detailed for patients diagnosed with hypermobile Ehlers Danlos Syndrome (hEDS, first column), Hypermobility Spectrum Disorder (HSD, second column), for all the patients combined (hEDS and HSD, third column in grey), and for age and gender matched local healthy controls (last column). BPI=Brief Pain Inventory; CSS16=Clinical Somatosensory Scale; SFN-SIQ=Small Fiber Neuropathy Symptoms Inventory Questionnaire; WPI=Widespread Pain Index; SS=Symptom Severity; QOL=Quality of Life from WHO-Bref; DN4=Douleur Neuropathique 4; HADS=Hospital Anxiety and Depression Scale; ***= p<0.001, differences between all patients and healthy controls. There were no significant differences between hEDS and HDS for these scores.

| Parameters |  | hEDS patients<br>(n=56) | HSD patients<br>(n=23) | All patients<br>(n=79) | Healthy controls<br>(n=23) |
| --- | --- | --- | --- | --- | --- |
| Age (years) | mean (SD) | 39.3 (11.6) | 40.2 (11.6) | 39.5 (11.5) | 39.5 (11.6) |
| Sex, n (%) |  |  |  |  |  |
|  | Male | 3 (5%) | 4 (17%) | 7 (9%) | 3 (13%) |
|  | Female | 53 (95%) | 19 (83%) | 72 (91%) | 20 (87%) |
| BPI Pain Severity | mean (SD)<br>n | 5.8 (1.7)<br>54 | 5.2 (2.1)<br>22 | 5.7 (1.9)<br>76 |  |
| BPI Pain Interference | mean (SD)<br>n | 6.0 (2.3)<br>53 | 4.8 (2.5)<br>22 | 5.7 (2.4)<br>75 |  |
| CSS-16 | mean (SD)<br>n | 39.6 (13.6)<br>47 | 33.9 (12.6)<br>18 | 38.0 (13.5)<br>65 |  |
| SFN-SIQ | mean (SD)<br>n | 18.8 (9.4)<br>51 | 17.7 (8.6)<br>21 | 18.5 (9.2)<br>72 | 2.3 (2.3) ***<br>23 |
| WPI | mean (SD)<br>n | 11.1 (4.2)<br>55 | 11.6 (3.9)<br>22 | 11.3 (4.1)<br>75 | 1 (0.9) ***<br>23 |
| SS | mean (SD)<br>n | 9.6 (2.2)<br>55 | 8.9 (2.2)<br>22 | 9.4 (2.2)<br>75 | 2.1 (1.1) ***<br>23 |
| QOL physical health | mean (SD)<br>n | 27.6 (15.7)<br>53 | 34.9 (21.6)<br>21 | 29.7 (17.7)<br>74 |  |
| QOL psychological health | mean (SD)<br>n | 51.4 (21.4)<br>53 | 60.2 (17.2)<br>21 | 53.9 (20.6)<br>74 |  |
| QOL social relationships | mean (SD)<br>n | 54.7 (23.4)<br>53 | 61.4 (20.6)<br>21 | 56.6 (22.7)<br>74 |  |
| QOL environment | mean (SD)<br>n | 57.5 (18.6)<br>53 | 66.1 (12.0)<br>21 | 59.9 (17.4)<br>74 |  |
| DN4 | mean (SD)<br>% of positivity (>3)<br>n | 4.5 (1.8)<br>72%<br>53 | 4.3 (1.9)<br>77%<br>22 | 4.4 (1.8)<br>73%<br>75 |  |
| HADS-Anxiety | mean (SD)<br>% of positivity (≥8)<br>n | 10.4 (4.0)<br>75%<br>53 | 10.5 (4.1)<br>77%<br>22 | 10.5 (4.0)<br>76%<br>75 |  |
| HADS-Depression | mean (SD)<br>% of positivity (≥8)<br>n | 8.3 (4.0)<br>47%<br>53 | 6.1 (3.8)<br>27%<br>22 | 7.7 (4.1)<br>48%<br>75 |  |
| Pain catastrophizing | mean (SD)<br>% of positivity (≥20)<br>n | 24.9 (12.6)<br>65%<br>52 | 21.2 (10.4)<br>50%<br>22 | 23.8 (12.0)<br>61%<br>69 |  |
| Kinesiophobia | mean (SD)<br>% of positivity (≥40)<br>n | 43.0 (11.5)<br>70%<br>53 | 37.8 (12.5)<br>41%<br>22 | 41.5 (12.0)<br>61%<br>75 |  |

The healthy controls filled the SFN-SIQ, the WPI and the SS. The other questionnaires were not collected, as they have clear clinical cut-offs or data from large reference populations.

### Pain body map analysis

Patients’ paper body maps from the BPI were scanned; the numerical images were processed using a custom-made program coded in Python, inspired by prior work [29]. Each pain drawing was superimposed to a template divided into 56 body segments. The merged image (drawing + segmentation) was evaluated by two researchers (A.F., M.V) blinded to the identity of the patient. They classified as active each body segment that was filled in some way (i.e. colored, containing a cross, or circled). This allowed to determine the pattern of active segments for each patient. Then, the percentage of patients that reported pain in each body area was calculated and represented on a heat-map [30].

### Quantitative sensory testing

Quantitative sensory testing (QST) was performed following the standardized protocol of the German Research Network Neuropathic Pain (DFNS) [31] by one experimenter (A.F.). After a log transformation of the raw data to follow a normal distribution, a z-score sensory profile was calculated using the formula: z-score=(value of the patient – mean value of published controls)/standard deviation of published controls).

Negative z-scores indicate loss of function and positive z-scores indicate a gain of function. For individual clinical assessment, each patient’s QST values were compared with the corresponding age and gender reference values from the literature [32]. In addition to individual assessments, group comparisons were performed between hEDS/HSD patients and an age- and gender-matched control group, locally recruited and tested in the same experimental conditions. This internal control was added to ensure the observed differences from the published controls could be attributed to patients’ specificities and not differences in experimental setting.

The QST parameters were acquired in the following order: cold detection threshold (CDT), warm detection threshold (WDT), ability to detect temperature changes (thermal sensory limen, TSL), the number of paradoxical heat sensations during TSL (PHS), cold pain threshold (CPT), heat pain threshold (HPT) assessed using TSA **II** Peltier thermode (Medoc, Israel). Mechanical detection threshold (MDT) was assessed using Von Frey filaments, mechanical pain threshold (pin/prick thresholds, MPT) and mechanical pain sensitivity (MPS) were assessed using pinpricks, dynamic mechanical allodynia (DMA) using a brush, cotton wool and a Q-tip, wind-up ratio represents the pain summation to repetitive pinprick stimuli (WUR), pressure pain threshold (PPT) was assessed using a calibrated algometer (Wagner Instruments, USA), and vibration detection threshold (VDT) using a tuning fork.The testing was performed on a hand and a foot, choosing the anamnestically more affected side except for 3 patients who were tested either on both hands (N=2) or both feet (N=1). For these 3 patients, the data of the most affected limb were considered for analysis. Thermal modalities were not assessed in 1 patient who could not tolerate the thermode due to reported electrosensitivity. CPT, HPT and MPS were not assessed in 1 other patient due to allodynia/hyperalgesia. Missing data were not replaced and the N for each modality is reported on Figure 4.

### Skin biopsies

Samples were collected at the Lausanne University Pain center and analyzed at Bern University Hospital (Inselspital) as part of routine clinical care. A skin punch biopsy (3mm; device by Stiefel, Germany) was performed under local anesthesia with subcutaneous lidocaine and a topical cold spray (Sintetica, Switzerland) to ensure comfort despite self-reported local anesthetic resistance in many patients. The skin sample was fixed in ready-to-use 4% paraformaldehyde (Biosystems, Switzerland) at 4°C for 1h and stored into a freshly prepared phosphate buffer until processed. The intraepidermal nerve fiber density count took place at the Neuromorphological laboratory of Bern University Hospital (Inselspital). As part of their standard procedure, 50μm cryosections were immunoreacted with antibodies against protein-gene product 9.5 (PGP9.5). PGP9.5-positive nerves were quantified visually using fluorescent microscope. The Inselspital-University of Bern Pathology institute method is based on [33]’s work and a count of IENFD >100/mm^2^ is considered normal.

### Small fiber neuropathy definition

Although there is not yet a gold standard for the diagnosis of SFN, based on recent recommendations [15, 16], in this cohort of patients with compatible symptoms, we considered there was a SFN if both the structure (=reduced IENFD) and the function (=decreased thermal detection at the hand or the foot on QST) were altered compared to normative values. SFN was excluded if both IENFD and QST were normal. Possible SFN was defined as only one abnormal test (IENFDor QST).

### Statistical analysis

All the analyses were performed using SPSS Statistics 27 (IBM, Germany). Demographic parameters were compared using unpaired t-tests. For individual patients’ QST analysis, Z scores above 1.96 (gain of function) or below -1.96 (loss of function) were considered as abnormal based on DFNS standards [32]. T-tests were performed to compare normally distributed z-scores between patients and the local control group for each modality. T-tests were used to perform group comparisons (hEDS vs HSD; patients vs controls, SFN vs no SFN). Bonferroni correction for multiple comparisons was applied with a reporting of adjusted p values. A Mann Whitney test was used to compare the number of patients reporting pain in different body areas (Body map data) between diagnostic categories (SFN, possible SFN, and no SFN). Pearson correlations were calculated between pain intensity, IENFD, Z-scores of QST thermal modalities, pain intensity (BPI) and CSS16.

## Results

### Demographic data and symptoms description

Demographic data for all the patients are detailed in Table 1. Of the 79 patients included in the analyses, 56 fulfilled hEDS diagnostic criteria (71%). The 23 HSD patients failed hEDS criteria because of missing a positive family history (N=10), systemic symptoms (N=3) or both (N=10). There were no significant differences in any of the symptom scales (CSS, BPI, WPI, SS, SFN-SIQ, DN4), quality of life nor psychological health between hEDS and HSD patients. All the patients reported pain (BPI-PS M=5.7, SD=1.9) interfering with their daily life (BPI-PI M=5.7, SD=2.4). The mean score for Widespread Pain Index was 11.3/19 (SD=4.1). Sixty of the 79 patients drew their pain on the body map, with the pain distribution reported in Figure 5C. On average, 58% of the body zones were considered as painful. Anxiety (76%), depression (48%), catastrophizing (61%) and kinesiophobia (61%) were frequently observed(Table 1).

### Small fiber neuropathy symptoms inventory questionnaire (SFN-SIQ)

All the patients reported both autonomic and sensory symptoms compatible with small nerve fiber impairment. The SFN-SIQ score was significantly higher in the hEDS/HSD patients group (M=18.5, SD=9.2) than in the local healthy control group (M=2.3, SD=2.3), t(90)=-9.5, p<0.001 (Figure 3).

**Figure 3.**
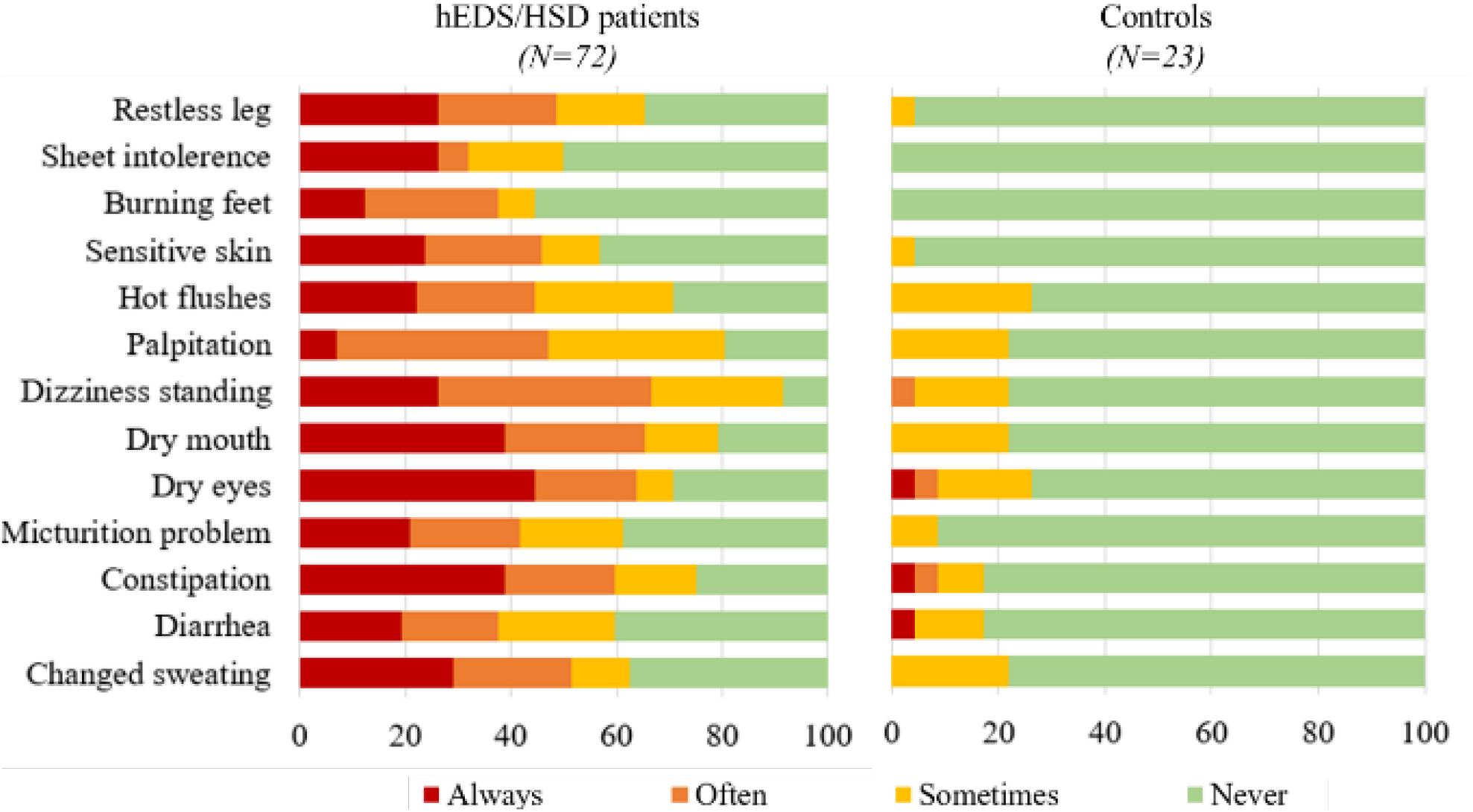
Frequency of small fiber neuropathy symptoms. Symptoms were reportedby hEDS/HSD patients and by the local matched sample of healthy controls using the Small Fiber Neuropathy Symptoms Inventory Questionnaire (SFN-SIQ).

### Quantitative sensory testing (QST)

Individual sensory profiles are represented in Figure 4. There were no significant differences in sensory profiles between patients diagnosed with hEDS and HSD, justifying the aggregation of the two diagnostic groups. Sensory loss of function of small fibers (i.e., any decrease in thermal detection: CDT, WDT or TSL on either hand or foot) was observed in 55/79 hEDS/HSD patients (70%) when compared to published normative data [32]. Specifically, more than one third of the patients displayed cold and/or for warm hypoesthesia (tables of Figure 4 A and B). Patients also presented hypodetection of temperature changes (TSL, hand: 46%; foot: 37%), hyperalgesia to muscle pressure (PPT, hand: 32%; foot: 14%) as well as hypoesthesia for light touch (MDT, hand: 42%; foot: 32%).

**Figure 4.**
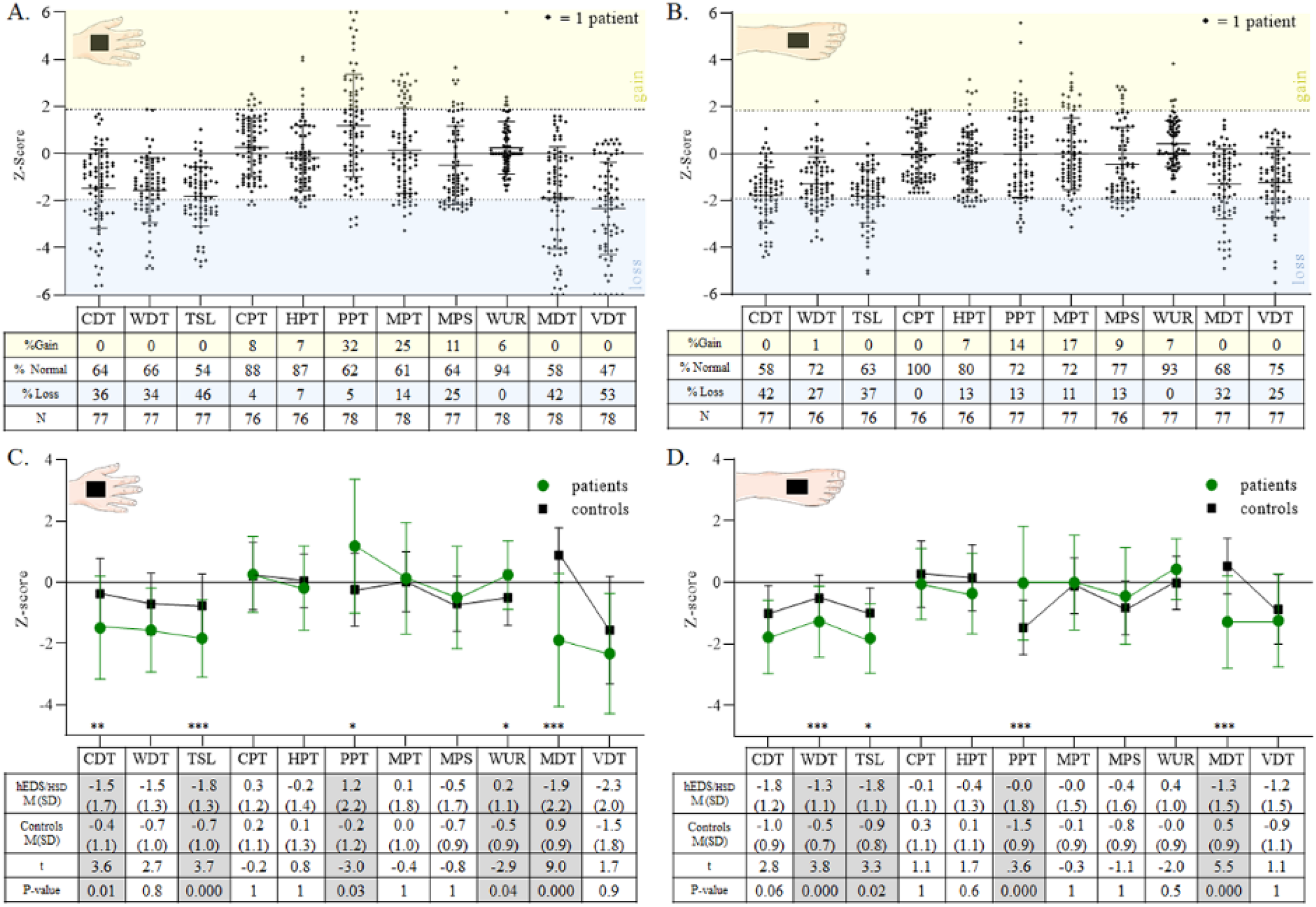
Quantitative Sensory Testing profile assessed on the most affected hand (A and C) and foot (B and D). Patients’ individual results are presented on the upper panels (A, B). Z-score beyond 1.96 (in the yellow band) correspond to a sensory gain of function and below -1.96 (in the blue band) to a loss of function, in comparison to published normative data. The N and the distribution between gain, loss or normal sensory function are presented for each modality in the tables below panels A and B. Group comparison between patients and the local matched sample of healthy controls are presented in the lower panels (C, D). The tables below panels C and D detail the mean Z-scores and standard deviations for each modality and the statistics: t and p-values, adjusted for multiple comparisons. Statistically significant differences are highlighted in grey. CDT=cold detection threshold. WDT=warmdetection threshold. TSL=thermal sensory limen. CPT=cold pain threshold. HPT=heat pain threshold. PPT=pressure pain threshold. MPT=mechanical pain threshold. MPS=mechanical pain sensitivity. WUR=wind-up ratio. MDT=mechanical detection threshold. VDT=vibration detection threshold. *** p<0.001, **p<0.01, *p<0.05.

When compared to the local matched controls, patients had significantly increased detection thresholds for cold, warm and temperature changes on both the hand and the foot (see Figure 3, panel C and D). These alterations are compatible with functional impairment of small fibers. In addition, patients displayed an increased wind-up ratio (WUR) as well as a decreased detection of light touch (MDT). Paradoxical heat sensations were observed in 6 patients for the hand (8%) and in 34 patients for the foot (45%) whereas none of the local controls experienced such sensation on the hand, and only two on the foot (9%). Dynamic mechanical allodynia was measured in 9 patients (12%) for the hand and in 11 patients for the foot (14%) but not in controls.

### Intraepidermal Nerve Fiber Density(IENFD)

There was no difference in IENFD between hEDS (M=77.4, SD=42.5) and HSD (M=62.4, SD=45.5), t(67)=0.8 p=0.4, justifying also a joint analysis. Small nerve fiber density was assessed in 69 of the 79 patients (48/56 hEDS and 21/23 HSD patients). IENFD ranged from 0 to 212 small nerve fibers by mm^2^ (M=74.7, SD=43.3). A reduced IENFD was described in 54 patients (78.2%) (Figure 5, panel A).

**Figure 5.**
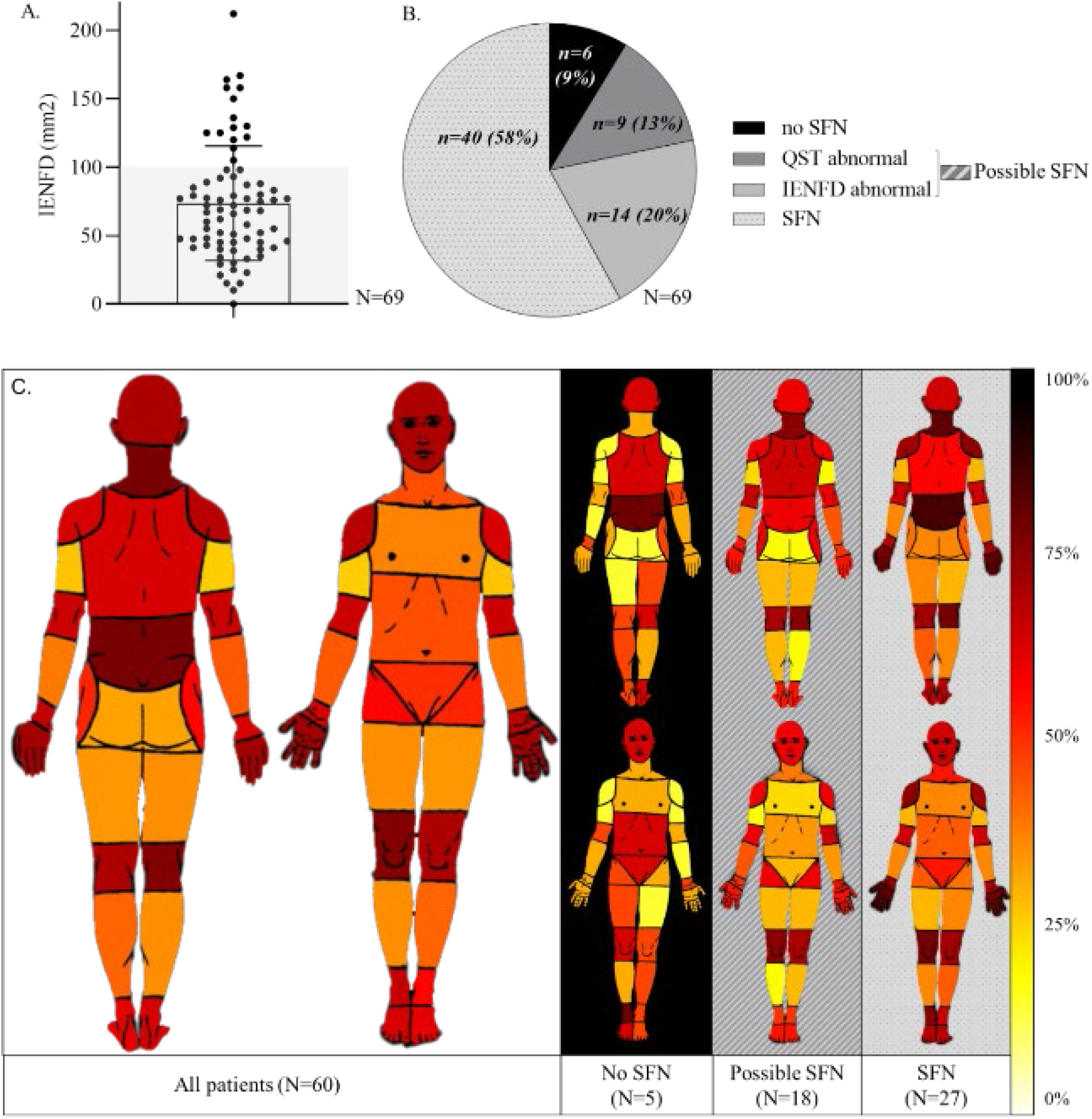
Intraepidermal nerve fiber density (IENFD) (A); subdivision of the hEDS/HSD population according to the likelihood of small fiber neuropathy based on QST and IENFD (B); pain distribution heat map (C). The left part of panel C represents the frequency of pain localization of hEDS/HSD patients as reported on pain drawings; the right panel shows the body map representations in sub-groups of patients based on their SFN likelihood. The color gradient represents the percentage of patients reporting pain in each body area from light yellow (0%) to dark red (100%). IENFD=intraepidermal nerve fiber density. QST=Quantitative Sensory Testing. SFN=Small Fiber Neuropathy with both QST and IENFD abnormal. No SFN=both QST and IENFD normal. Possible SFN=either QST or IENFD abnormal.

### Correlations between IENFD and QST

There was no correlation between IENFD and Z-scores of thermal QST modalities. Furthermore, no correlation was shown between these small fiber integrity outcomes and symptom severity questionnaires (pain intensity and interference, CSS16, SFN-SIQ).

### Small fiber neuropathy (SFN)

SFN defined as a decrease of both IENFD and thermal detection, was confirmed in 40/69 patients (58%). Possible SFN was detected in 23/69 patients. SFN was excluded in 6 patients (Figure 5 B). Among the 10 patients who did not undergo skin biopsy for IENFD count, 6 displayed functional abnormalities (QST).

Fifty out of the 79 patients (63%) underwent an ENMG, which was normal in 40 (80%). As a reminder, this procedure was not performed at our center, and hence, some patients did not comply with the recommendations to consult a neurologist. Among the 10 abnormal ENMGs, 7 revealed a carpal tunnel syndrome, 1 a tarsal tunnel syndrome, 1 a meralgia paresthetica and 1 a distal polyneuropathy. The patient with a polyneuropathy was kept in the SFN group for analysis according to the chosen definition based on structural and functional criteria.

### Differences between patients with and without SFN

The distribution of the pain localization for the hEDS/HSD population based on the body maps (filled by 60/79) is presented in Figure 5C (left panel). A sub-group analysis according to the likelihood of SFN is shown on the right panel of Figure 5C. The amount of patients reporting pain in the hand significantly differed among these categories (SFN, possible SFN, no SFN; H(2)=8.6, p=0.01), with a post hoc Bonferroni test showing a significant difference between patients with and without SFN (adjusted p=0.03). Patients with and without SFN showed no significant differences in terms of BPI, CSS16, WPI, SS, psychological scores nor SFN-SIQ.

## Discussion

We observed a small nerve fiber dysfunction based on quantitative sensory testing (QST) as well as a structural deficit based on intraepidermal nerve fiber density (IENFD) in a large cohort (N=79) of patients suffering from hEDS/HSD. These patients were referred to an academic pain center for a small fiber pathology assessment based on anamnestic complaints (burning pain, allodynia or dysautonomia). SFN was definite (both abnormal structure and function) in 40/69 patients (58%), possible (one abnormal measure) in 23/69 patients (33%) and excluded (both normal) in only 6/69 patients (9%). These results are consistent with the first description of small nerve fiber pathology based on IENFD counts in a smaller cohort of hEDS patients (N=20) [20].

Interestingly, previous QST studies did not reveal a decreased thermal detection [34, 35] in small cohorts (27 and 22 patients) of hEDS/HSD patients complaining of pain with a negative DN4 score. Yet, the presence of neuropathic pain characteristics has been reported frequently in hEDS/HSD [11, 12, 36]. In our work, 50% of a hEDS/HSD cohort were referred to our center for neuropathic symptoms. Thermal hypoesthesia was previously shown, yet not measured according to the standard DFNS protocol in a population of 37 hEDS patients with neuropathic pain [11]. Hence, our data confirmed the latter results with a standardized QST methodology, in a larger cohort of hEDS/HSD patients with neuropathic pain symptoms.

Even though QST is a widely used test for functional evaluation of SFN, it is a subjective test with potential inter-observer variability. In our sample, 53% of patients had hypopallesthesia when compared to published normative data [32]. Yet, when comparing patients to the local control group, there was no significant difference in VDT. This highlights the usefulness of local control groups for QST, as recommended by others [37].

Furthermore, our study, bringing together functional and structural assessments with both QST and IENFD abnormalities in 58% of patients, expands upon prior single-method findings. Nevertheless, there is no significant correlation between IENFD and thermal QST modalities indicating a discrepancy between functional and morphological assessments. Yet the correlation between QST and IENF density remains controversial [38] and several methodological considerations apply [37]. First the PGP9.5 marker used for small fiber staining cannot distinguish between subtypes of small fibers and their specific functions [39]. Additionally no information is obtained about the receptors present and active at their membrane [40]. Finally, the nerve fiber excitability is not assessed through such histological staining, leading to a possible normal count of nonfunctional fibers. A supplementary hypothesis proposes compensatory spinal and supraspinal mechanisms leading to normal thermal detection despite decreased fiber density [41].

Interestingly, on average, hEDS/HSD patients presented increased light touch detection thresholds (Von Frey filaments, considered to reflect large fiber function), despite otherwise normal large fiber testing (NCS, VDT). This impaired tactile detection could be caused by a subgroup of tactile C-fibers afferents responsible for the conduction of pleasant touch [42]. An alteration in this subgroup of mechanosensitive C-fibers might also explain the loss of function in light touch detection observed in our sample. Touch modulates pain signals, therefore a decreased information from touch-receptors could lead to a lack of pain alleviation [43]. These sensory deficits, as well as a lack of proprioception [44] could lead to an imbalance of inputs in hEDS/HSD toward increased nociception.

The pain drawing analysis revealed the hands were more frequently painful in patients with SFN but none of the standardized questionnaires revealed differences between patients with or without SFN. The SFN-SIQ score while significantly different between hEDS/HSD patients and controls, failed to discriminate for the presence of SFN.

Despite the few differences in phenotype between hEDS/HSD patients with and without SFN, adding this diagnosis seems important since patients with SFN defined by histological findings respond better to neuropathic pain medication [45].

There was no difference in pain intensity between patients with and without SFN and no correlation between SFN outcomes (IENFD and QST thermal modalities) and pain severity. Three reasons could explain this. First the missing fibers may not be the ones ‘causing’ pain, they could be C-touch fibers as discussed above. Second, morphologically normal nerve fibers may be sensitized and show increased activity [46, 47]. Finally, pain in hEDS/HSD can have other contributing factors than nerve damage.

The large proportion of our patients displaying small nerve fiber abnormalities underscores the possible contribution of the peripheral nervous system to hEDS/HSD symptoms. Interestingly, unmyelinated small fibers (sympathetic and sensory axons) contribute to 80% of joint innervation and to other components of the musculoskeletal system such as the tendons [48] [49]. Afferent innervation also affects several aspects of muscle development, including muscle spindle formation and maintenance [50]. A loss of sensory innervation impairs healing of bone fractures and may contribute to chronic musculoskeletal diseases [51]. Hence, peripheral nerves contribute to musculoskeletal tissue development and homeostasis [51]. Given the observed autonomic symptoms and the sensory deficits, we can hypothesize that in a subgroup of patients, the small nerve pathology could participate in the hypermobile pathophysiology. It could be a causal factor during development, or a secondary factor leading to impaired healing from repetitive trauma.

Nevertheless, the pain experience in hEDS/HSD is likely to be multifactorial. There is a nociceptive contribution with joint instabilities resulting in repetitive dislocations, a muscular component or periarticular inflammation [52]. In our cohort, large joints were amongst the most frequently designated painful areas on the pain drawings (Figure 4 C). Central sensitization is a likely feature, given a previous demonstration of hyperalgesia in non-painful areas and an increased wind-up ratio compared to healthy controls (yet not reaching the DFNS cut-off) [34, 35] as well as a deficit of descending inhibitory pain control using a conditioned pain modulation protocol [34]. We observed an increased wind-up ratio following DFNS criteria on average between patients and local controls. However, only 7/79 patients showed a significant Z-score elevation (>1.96) compared to the published normative values [32]. We observed hyperalgesia to muscular pressure and dynamic mechanical allodynia but no thermal hyperalgesia. Hence, in our cohort, there is evidence for diverse pain contributors including peripheral neuropathic, nociceptive musculoskeletal, and nociplastic. The proportion of each of these factors appears to be different depending on publications, which is not surprising given the heterogeneity of the hEDS/HSD population and various recruitment criteria.

In our cohort, there were no significant differences on any of the measures between patients fulfilling 2017 hEDS or HSD criteria. These results further question whether hEDS and HSD, while separated by the 2017 diagnostic criteria, are two different entities [5, 53]. Additionally, none of the classification systems categorizing hypermobility syndromes, whether by Villefranche, Brighton or Malfait 2017, appears to stratify successfully patients according to severity [53]. Indeed, many experts in the field now consider a single phenotype termed hEDS/HSD [5, 53].

Dysautonomia has been prospectively investigated in hEDS in the cardiovascular, sudomotor and gastrointestinal systems. Studies revealed an over-activity of the resting parasympathetic tone and a decreased sympathetic reactivity to stimuli [54]. Interestingly, cardiovascular dysautonomia and a range of other functional symptoms commonly reported in hEDS/HSD are also considered typical manifestations of SFN [55].The development of dysautonomia of potential neurogenic origin could represent a determining factor for the difference between a simple and a more severe form of hEDS/HSD [53]. Our cohort was not tested for dysautonomia except for the SFN-SIQ, and further work will follow this direction.

Even though this research is promising, some limitations need to be addressed. First, the retrospective design entails some significant limitations: patients were referred to the pain center because of symptoms compatible with SFN (dysesthesia or dysautonomia). Hence, the described cohort is reflecting a small but significant proportion (50%) of a full population of hEDS/HSD patients (as described in [5]). Assessments of an entire cohort could determine the SFN prevalence in a general population of hEDS/HSD, including those who are asymptomatic. Also due to the retrospective design, some NCSs were missing leading to a limitation in the diagnosis of SFN based on recent, although still debated recommendations [14]. Nevertheless, the potential co-occurrence of a large fiber dysfunction could suggest a mixed contribution, but does not invalidate the small fiber pathology.

Furthermore, due to the delayed availability of skin biopsy readings, there was a lag up to 3 years between QST and IENFD assessments, raising the question of the natural evolution of SFN and neuropathic pain in the hypermobile syndrome.

In conclusion, clinicians should assess small fiber pathology in hEDS/HSD patients for sensory and autonomic impact. The presence of small fiber pathology could be used to better stratify the heterogeneous hEDS/HSD population in terms of severity and disease extent. Our results also open new avenues for the pathophysiological research, since the hypermobile form is the only EDS syndrome without a known genetic cause. The search should possibly be broadened to a neurological origin. A better understanding of the pathophysiology of the hypermobility syndromes could facilitate more efficient symptom management. Furthermore, the co-occurrence with SFN, a neurological disorder that can be diagnosed by objective measures, might improve the recognition of the complex hEDS/HSD pathology.

## Data Availability

All data produced are available on Zenodo
DOI: 10.5281/zenodo.6123805

https://zenodo.org/record/6123805#.Yg5ffDHMKUk

## Acknowledgments

We are grateful to Prof. Isabelle Décosterd, chair of the Pain Center, Lausanne University Hospital, for her support and the nurturing research environment she has created. We also thank Prof. Charles Benaim, Service of rheumatology, Lausanne University Hospital, for his kind collaboration around the hEDS/HSD cohort. We are grateful to Dr Nicole Kamber and Mrs Martina Schobesberger from the Inselspital Neuromorphology Laboratory, Bern University Hospital, for the harmonious clinical collaboration.

